# A novel Aβ PET scoring system for predicting the response of Alzheimer’s disease to lymphatic-venous anastomosis

**DOI:** 10.64898/2026.07.08.26357543

**Authors:** Jian Liu, Pengfei Li, Zhenyu Luo, Chen Li, Xiaoguang Du, Nana Wang, Haijun Li, Tingzhong Wang, Xiaoya Feng

## Abstract

**Objective:** Deep cervical lymphatic-venous anastomosis (LVA) has shown promise in treating Alzheimer’s disease (AD), yet no preoperative tool exists to identify potential responders. We developed and evaluated a novel Aβ PET-based scoring system that quantifies regional amyloid burden according to anatomical proximity to the meningeal lymphatic vessels (MLVs) to predict treatment response.

**Methods:** We retrospectively enrolled 58 AD patients who had undergone upper cervical LVA. Eleven regions of interest (ROIs) adjacent to the superior sagittal and straight sinuses were scored based on anatomical proximity to MLVs (higher = closer) and functional relevance to AD (functional score = 1 for AD-related ROIs), yielding a regional assigned score (RAS). Standardized uptake value ratios (SUVRs) were obtained for each ROI. The total SUVR (S_total_) was calculated as Σ(SUVR × RAS) over all ROIs, and S_4+5_ was defined as the same sum restricted to ROIs with RAS 4 or 5. These scores, along with baseline demographic characteristics, were evaluated for their ability to predict treatment response using LASSO-logistic regression and receiver operating characteristic (ROC) curve analysis.

**Results:** Forty-one patients (70.7%) were responders. At baseline, responders had significantly higher SUVR of the associative visual cortex (S_AVC_) (1.68±0.26 vs. 1.53±0.12, P=0.0394) and higher S_4+5_ (32.69±4.45 vs. 30.14±3.07, P=0.0358) than non□responders. In univariate analysis, S_4+5_ was the only significant predictor (OR=1.183, 95% CI: 1.005–1.391, P=0.0433); S_AVC_ was borderline significant (OR=16.654, 95% CI: 0.999–277.63, P=0.0501), while SUVR of the posterior cingulate cortex (S_PCC_) and Mini□Mental State Examination (MMSE) showed only weak trends (P=0.0714 and P=0.0889, respectively). In the multivariable model, MMSE was independently associated with treatment response (adjusted OR = 1.43, 95% CI: 1.06–1.93, P = 0.022); with S_PCC_ and SUVR of the superior parietal cortex (S_sPL_) reaching marginal significance (P=0.055 and P=0.051, respectively). The apparent AUC was 0.920, decreasing to a Bootstrap-corrected AUC of 0.780 (95% CI: 0.708–0.884) after optimism correction (optimism = 0.139). The Brier score was 0.097. The covariates-only model yielded a corrected AUC of only 0.574, confirming the incremental value of PET DOI data.

**Conclusion:** This exploratory study introduces a novel Aβ PET scoring system grounded in MLV anatomy that, combined with baseline MMSE, demonstrates modest predictive potential for LVA response in AD. The findings warrant validation in larger, multicenter cohorts.

## Introduction

Alzheimer’s disease (AD) is the leading cause of dementia, characterized neuropathologically by extracellular amyloid□β (Aβ) plaques, intraneuronal tau tangles, and progressive synaptic and neuronal loss. Despite decades of drug discovery, currently approved therapies offer only modest symptomatic relief or a marginal slowing of early cognitive decline, without halting disease progression [1–4]. Deep cervical lymphatic-venous anastomosis (LVA), a novel microsurgical strategy first reported in 2018 [5], aims to enhance cerebral Aβ clearance by facilitating drainage into the peripheral lymphatic system. However, LVA response rates vary considerably across centers (60–84.2%) [6–8], and no preoperative predictor exists to identify likely responders.

The anatomical basis for LVA lies in the meningeal lymphatic vessels (MLVs), which run along the superior sagittal sinus (SSS) and straight sinus (SS) and serve as the primary conduit for neurotoxic proteins drainage to cervical lymph nodes [9,10]. As the most important diagnostic tool for AD, Aβ positron emission tomography (PET) enables in vivo quantification of fibrillar amyloid deposition [11], but conventional global metrics (e.g., Centiloid) do not account for regional heterogeneity or anatomical relationships to MLVs. The most widely used semi□quantitative metric is the standardized uptake value ratio (SUVR), which normalizes tracer retention in a target region to a reference area with low fibrillar amyloid content [12].

We hypothesized that the hydrodynamic efficacy of LVA depends on the spatial distribution of Aβ burden relative to MLV drainage pathways: regions closer to MLVs (i.e., adjacent to the SSS/SS) may benefit more from enhanced clearance, provided they harbor sufficient Aβ substrate. Building on this rationale, we developed a novel scoring system that integrates (1) anatomical proximity to MLVs, (2) functional relevance to AD pathophysiology, and (3) quantitative Aβ burden from PET. Using LASSO-regularized logistic regression, we then built and internally validated a predictive model for postoperative cognitive response to LVA.

## Methods

### Study Design and Participants

This retrospective study was approved by the institutional Ethics Committee. We enrolled 58 patients with AD who underwent bilateral upper cervical LVA at our center between January 2025 and June 2025. Inclusion criteria were: (1) age 50–85 years; (2) diagnosis of AD based on 2024 NIA-AA revised criteria (core 1 biomarkers positive confirmed by Aβ PET with or without CSF); (3) Mini-Mental State Examination (MMSE)≤ 18 and Clinical Dementia Rating (CDR) ≥1.

Exclusion criteria were: (1) Dementia not confirmed by Aβ PET; (2) MMSE > 19 or CDR<1; (3) Absence of complete clinical data. All patients provided informed consent.

### PET Acquisition and the Novel Scoring System

PET/CT scans were acquired on a United Imaging uMI Vista Pro scanner. Each patient received an intravenous bolus of ∼10 mCi (370 MBq) of ¹□F□Florbetaben (AV□1). A static 20□30 min PET acquisition (single bed position) was started 90 min post□injection. A low□dose CT (120 kV, 320 mAs) was performed for attenuation correction. PET data were reconstructed using TOF□OSEM (5 iterations, 20 subsets) with HD resolution recovery, applying corrections for random coincidences, normalization, attenuation, and head motion. A 6□mm Gaussian post□filter was used. Final images had a 256×256 matrix, 1.5□mm slice thickness, and 300□mm FOV. Images were fed into the NeuroQ (Syntermed) software, which automatically performed analysis following rigid registration to a standard brain database. The software automatically labeled brain regions of interest (ROIs) and calculated the SUVR for each ROI using the whole cerebellum as the reference region.

Given that human MLVs run parallel to the SSS and SS, each ROI received a regional assigned score (RAS) comprising an anatomical score (0.5–5, higher for closer proximity to SSS/SS) and a functional score (1 for ROIs linked to AD core symptoms) (Fig 1). Per NeuroQ protocol, a ROI with SUVR > 1.31 receives its RAS; otherwise, it contributes 0. The sum of RAS across all ROIs yielded total assigned scores (TAS). The total SUVR (S_total_) was calculated as Σ(SUVR × RAS) over all ROIs, and S_4+5_ as the same sum restricted to ROIs with RAS 4 or 5. At this point, all SUVR values are incorporated, regardless of the value.

**Figure 1.**
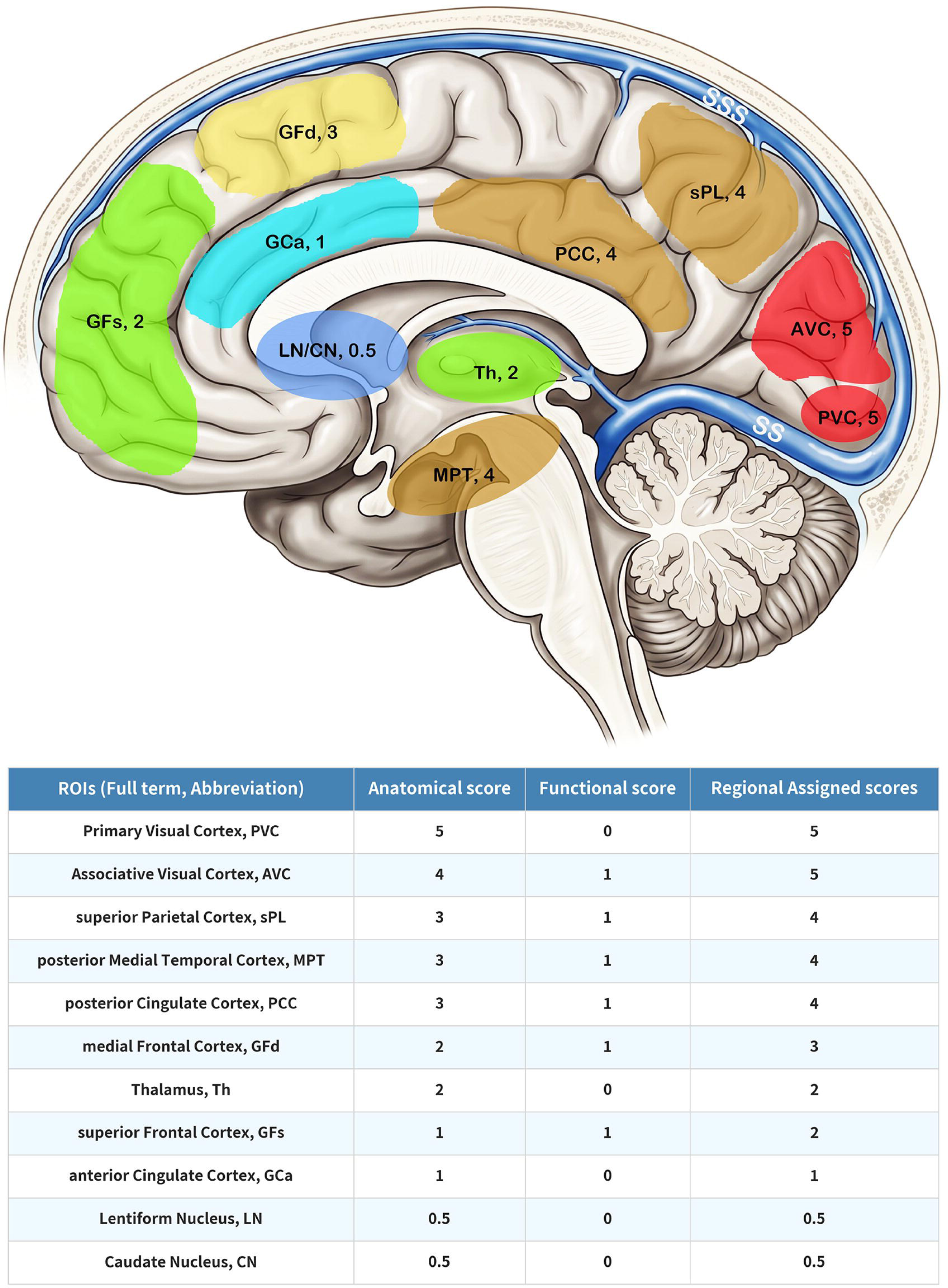
Design of the novel scoring system. Eleven ROIs adjacent to the SSS and SS are selected (top). The regional assigned score (RAS) for each ROI is the sum of its anatomical and functional scores (bottom). ROI: region of interest, SS: straight sinus, SSS: superior sagittal sinus.

### Surgical Procedure

Bilateral upper cervical LVA was performed under general anesthesia through a 5–6 cm incision along the anterior border of the sternocleidomastoid muscle. The internal jugular vein (IJV) and its primary tributaries were dissected and mobilized. The jugulodigastric node (JDN) was identified anterosuperior to the IJV (level IIA). Only the lower half of the JDN was exposed to avoid over-dissection that could damage the attached lymphatic vessels (Fig. 2A). The IJV was retracted anteriorly to expose the adipose tissue at level III. A U-shaped fat pad containing lymphoid tissue was dissected to create a cephalad-pedicled lymphatic flap (LF).

**Figure 2.**
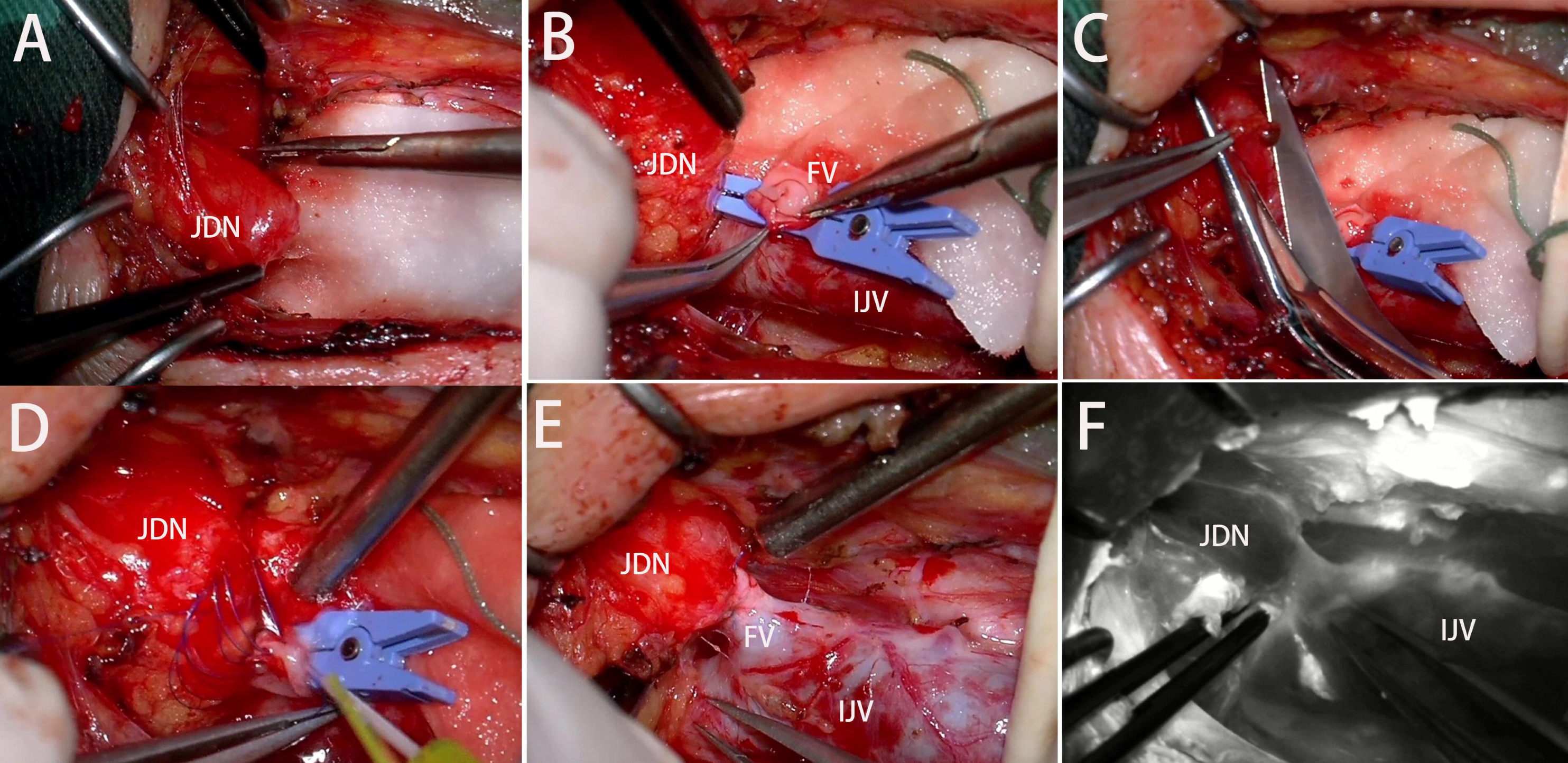
The surgical steps of LVA at level IIA. After dissecting the JDN (A), the FV is divided and a fish□mouth incision is made (B). The JDN is transected to create a cross□sectional stump (C), which is then anastomosed to the FV with the back wall sutured via the parachute technique (D). The JDN-FV end□to□end anastomosis is completed (E), and patency is finally verified by fluorescence angiography (F). FV: facial vein, IJV: internal jugular vein, JDN: jugulodigastric node.

At level IIA, the JDN was anastomosed end-to-end to the facial or lingual vein. The distal end of the facial or lingual vein was fashioned into a fish-mouth configuration (Fig. 2B). The JDN was transected to create a cut surface (Fig. 2C). The anastomosis was completed using 7-0 Prolene sutures. Because the JDN could be mobilized only to a limited extent and could not be flipped for convenient suturing, an intracorporeal parachute technique was employed for back-wall suturing (Fig. 2D). Before closure of the anterior wall, the anastomotic lumen was irrigated and filled with heparinized water (Fig. 2E). At level III, the same technique was used for end-to-side anastomosis of the LF to the IJV.

Anastomotic patency was evaluated using intraoperative indocyanine green fluorescence angiography. In general, when the recipient vessel is the facial vein or IJV, the fluorescent lymphatic fluid enters a large-caliber, high-flow vein and does not produce extensive fluorescence within the vein. Patency was confirmed only when gentle compression of the recipient vein with forceps elicited a smoke-like fluorescent flow across the anastomosis (Fig. 2F). Routine postoperative care was provided, and bilateral drains were removed within 48 hours.

### Data Acquisition

Baseline demographic characteristics, cerebrospinal fluid (CSF) AD biomarker results, and perioperative neuropsychological assessment data were retrieved from the electronic medical record system. CSF biomarkers were measured using enzyme-linked immunosorbent assay, with positivity defined as follows: Aβ42 ≤ 650 pg/mL, Aβ42/Aβ40 ratio < 0.05, P□Tau181 ≥ 58 pg/mL, and T□Tau ≥ 399 pg/mL.

Neuropsychological assessments were administered using standardized instruments, including the Mini□Mental State Examination (MMSE) and Montreal Cognitive Assessment (MoCA) for global cognition, the Clinical Dementia Rating Scale (CDR) for dementia staging, and the Activities of Daily Living (ADL) scale for functional status. On postoperative day 7 (before discharge), a patient was considered a responder if they achieved an improvement of ≥ 2 points on the MMSE or MoCA, or a reduction of ≥ 1 point on the CDR, or an improvement of ≥ 5 points on the ADL, and the improvement was also confirmed by caregiver report.

### Statistical Analysis

Normality was assessed using the Shapiro-Wilk test. Normally distributed continuous variables were compared by independent-samples t-test and expressed as mean ± SD; non-normal variables were compared by Mann-Whitney U test and expressed as median (IQR). Categorical variables were compared by χ² or Fisher’s exact test. Univariable logistic regression was performed for each predictor to estimate crude ORs.

Collinearity was assessed by variance inflation factor (VIF); VIF > 5 indicated moderate collinearity. LASSO-logistic regression with 10-fold cross-validation was used for variable selection. Continuous variables were Z-score standardized. The lambda.1se criterion was chosen for parsimony. Retained variables were entered into multivariable logistic regression (post-selection inference). Internal validation used 1000-bootstrap resampling to estimate optimism-corrected AUC and 95% CI. Model calibration was assessed by Brier score and Hosmer-Lemeshow test; clinical usefulness by decision curve analysis (DCA). A covariates-only model (without PET data) and a backward stepwise model (AIC-based) served as sensitivity analyses. All analyses were performed in Python 3.12; two-sided P < 0.05 was considered significant.

## Results

### Baseline Characteristics

Of 58 enrolled patients, 41 (70.7%) were responders and 17 (29.3%) were non-responders.

The mean age was 70.6 ± 8.2 years (range 56–85), with 21 males (36.2%) and 37 females (63.8%). Diagnostic information is summarized in Table 1. All patients were Aβ PET-positive. Responders showed significantly higher S_AVC_ (1.68±0.26 vs. 1.53±0.12, P=0.0394) and S_4+5_ (32.69±4.45 vs. 30.14±3.07, P=0.0358) than non-responders. Responders also exhibited higher MMSE scores (8 [2–11] vs. 5 [0–6], P=0.0865) and S_PCC_ (1.67±0.35 vs. 1.49±0.24, P=0.0604), both approaching statistical significance (Table 2).

**Table 1.** Diagnostic information.

| Category | Subcategory | Value |
| --- | --- | --- |
| **Positive Core 1 biomarkers** | Aβ PET + CSF Aβ42/40 + CSF p-tau181/Aβ42 + CSF t-tau/Aβ42 (4) | 20 (34.48%) |
| | Aβ PET + CSF p-tau181/Aβ42 + CSF t-tau/Aβ42 (3) | 27 (46.55%) |
| | Aβ PET only (1) | 11 (18.97%) |
| **Baseline neuropsychological scores** | MMSE | 6 (2-11) |
|  | MoCA | 3 (0-6) |
|  | CDR | 2 (2-3) |
|  | ADL | 70 (50-90) |
| **Integrated stage** | 5A | 41 (70.7%) |
|  | 6A | 17 (29.3%) |

**Table 2.**
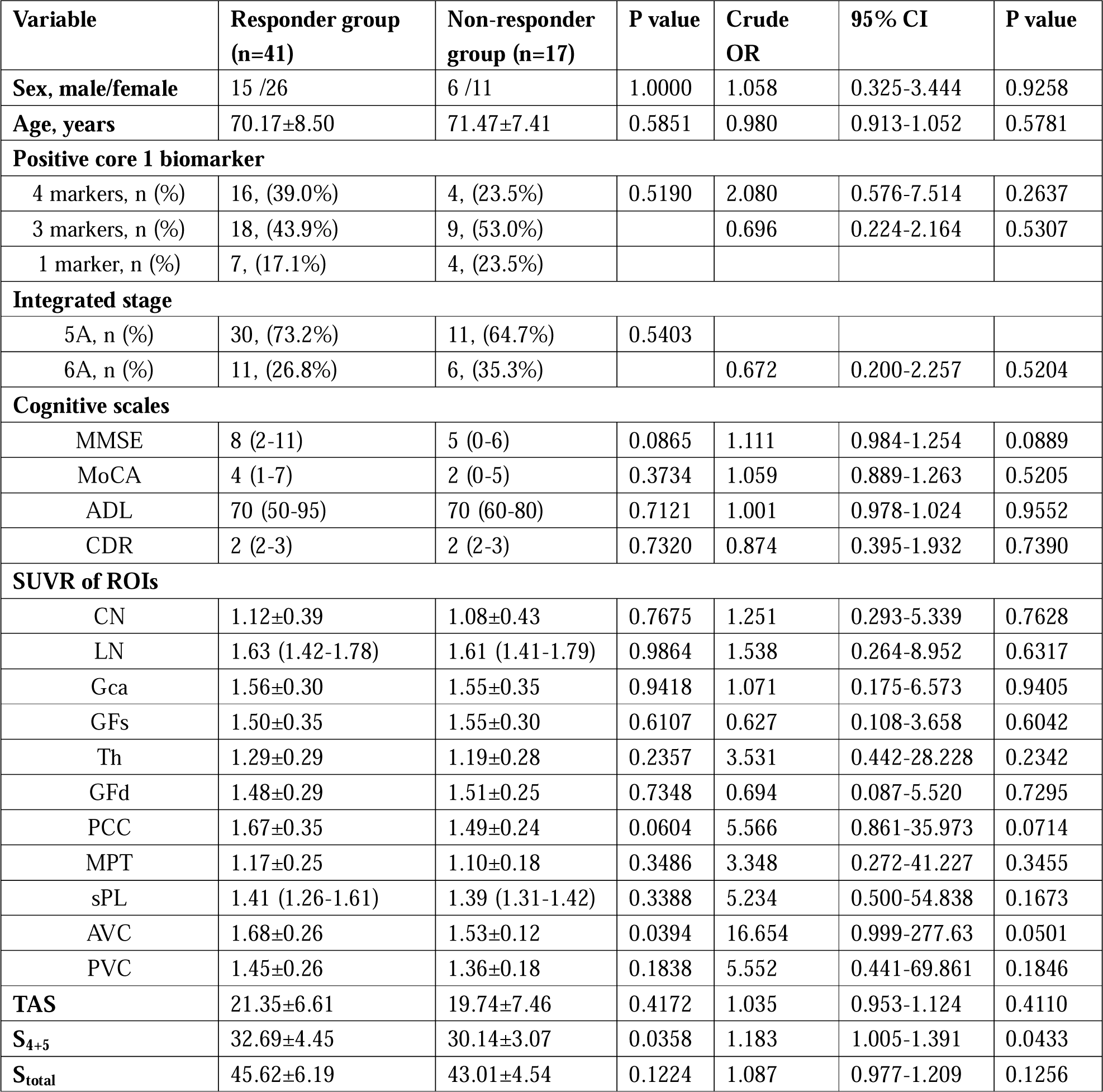
Intergroup Comparisons and Univariate Logistic Regression Analysis.

| Variable | Responder group<br>(n=41) | Non-responder<br>group (n=17) | P value | Crude<br>OR | 95% CI | P value |
| --- | --- | --- | --- | --- | --- | --- |
| Sex, male/female | 15 /26 | 6 /11 | 1.0000 | 1.058 | 0.325-3.444 | 0.9258 |
| Age, years | 70.17±8.50 | 71.47±7.41 | 0.5851 | 0.980 | 0.913-1.052 | 0.5781 |
| **Positive core 1 biomarker** |  |  |  |  |  |  |
| 4 markers, n (%) | 16, (39.0%) | 4, (23.5%) | 0.5190 | 2.080 | 0.576-7.514 | 0.2637 |
| 3 markers, n (%) | 18, (43.9%) | 9, (53.0%) |  | 0.696 | 0.224-2.164 | 0.5307 |
| 1 marker, n (%) | 7, (17.1%) | 4, (23.5%) |  |  |  |  |
| **Integrated stage** |  |  |  |  |  |  |
| 5A, n (%) | 30, (73.2%) | 11, (64.7%) | 0.5403 |  |  |  |
| 6A, n (%) | 11, (26.8%) | 6, (35.3%) |  | 0.672 | 0.200-2.257 | 0.5204 |
| **Cognitive scales** |  |  |  |  |  |  |
| MMSE | 8 (2-11) | 5 (0-6) | 0.0865 | 1.111 | 0.984-1.254 | 0.0889 |
| MoCA | 4 (1-7) | 2 (0-5) | 0.3734 | 1.059 | 0.889-1.263 | 0.5205 |
| ADL | 70 (50-95) | 70 (60-80) | 0.7121 | 1.001 | 0.978-1.024 | 0.9552 |
| CDR | 2 (2-3) | 2 (2-3) | 0.7320 | 0.874 | 0.395-1.932 | 0.7390 |
| **SUVr of ROIs** |  |  |  |  |  |  |
| CN | 1.12±0.39 | 1.08±0.43 | 0.7675 | 1.251 | 0.293-5.339 | 0.7628 |
| LN | 1.63 (1.42-1.78) | 1.61 (1.41-1.79) | 0.9864 | 1.538 | 0.264-8.952 | 0.6317 |
| Gca | 1.56±0.30 | 1.55±0.35 | 0.9418 | 1.071 | 0.175-6.573 | 0.9405 |
| GFs | 1.50±0.35 | 1.55±0.30 | 0.6107 | 0.627 | 0.108-3.658 | 0.6042 |
| Th | 1.29±0.29 | 1.19±0.28 | 0.2357 | 3.531 | 0.442-28.228 | 0.2342 |
| GFd | 1.48±0.29 | 1.51±0.25 | 0.7348 | 0.694 | 0.087-5.520 | 0.7295 |
| PCC | 1.67±0.35 | 1.49±0.24 | 0.0604 | 5.566 | 0.861-35.973 | 0.0714 |
| MPT | 1.17±0.25 | 1.10±0.18 | 0.3486 | 3.348 | 0.272-41.227 | 0.3455 |
| sPL | 1.41 (1.26-1.61) | 1.39 (1.31-1.42) | 0.3388 | 5.234 | 0.500-54.838 | 0.1673 |
| AVC | 1.68±0.26 | 1.53±0.12 | 0.0394 | 16.654 | 0.999-277.63 | 0.0501 |
| PVC | 1.45±0.26 | 1.36±0.18 | 0.1838 | 5.552 | 0.441-69.861 | 0.1846 |
| **TAS** | 21.35±6.61 | 19.74±7.46 | 0.4172 | 1.035 | 0.953-1.124 | 0.4110 |
| **S<sub>4+5</sub>** | 32.69±4.45 | 30.14±3.07 | 0.0358 | 1.183 | 1.005-1.391 | 0.0433 |
| **S<sub>total</sub>** | 45.62±6.19 | 43.01±4.54 | 0.1224 | 1.087 | 0.977-1.209 | 0.1256 |

In univariate logistic regression, S_4+5_ was the only significant predictor (OR=1.183, 95% CI: 1.005–1.391, P=0.0433), corresponding to approximately 18.3% increased odds per 1-point increment. S_AVC_ reached borderline significance (OR=16.654, 95% CI: 0.999–277.63, P=0.0501), whereas S_PCC_ (OR=5.566, 95% CI: 0.861–35.973, P=0.0714) and MMSE (OR=1.111, 95% CI: 0.984–1.254, P=0.0889) displayed only weak predictive trends (Table 2).

### LASSO Variable Selection and Multivariable Model

A LASSO□Logistic combined prediction model was constructed using the 11 DOI SUVRs as core predictors and other baseline variables as covariates. Collinearity diagnostics revealed no variance inflation factor (VIF) > 10, indicating acceptable multicollinearity. After Z□score standardization of the 11 SUVRs, 10□fold cross□validation LASSO regression was performed for variable selection. The lambda.1se criterion (λ = 0.756) was applied to favor a more parsimonious model, ultimately retaining 8 DOI SUVRs: GFs, Th, GFd, PCC, MPT, sPL, AVC, and PVC (Supplementary data 1). These 8 LASSO□selected indicators, together with 9 covariates, were entered into a multivariable logistic regression model for post□selection inference. After adjustment for covariates, S_PCC_ (adjusted OR=280.94, 95% CI: 0.89–88403.93, P=0.055) and S_sPL_ (adjusted OR=644.48, 95% CI: 0.98–423696.07, P=0.051) showed marginally significant positive associations, while MMSE remained significantly associated with treatment efficacy (adjusted OR=1.79, 95% CI: 1.09–2.95, P=0.022) (Figure 3).

**Figure 3.**
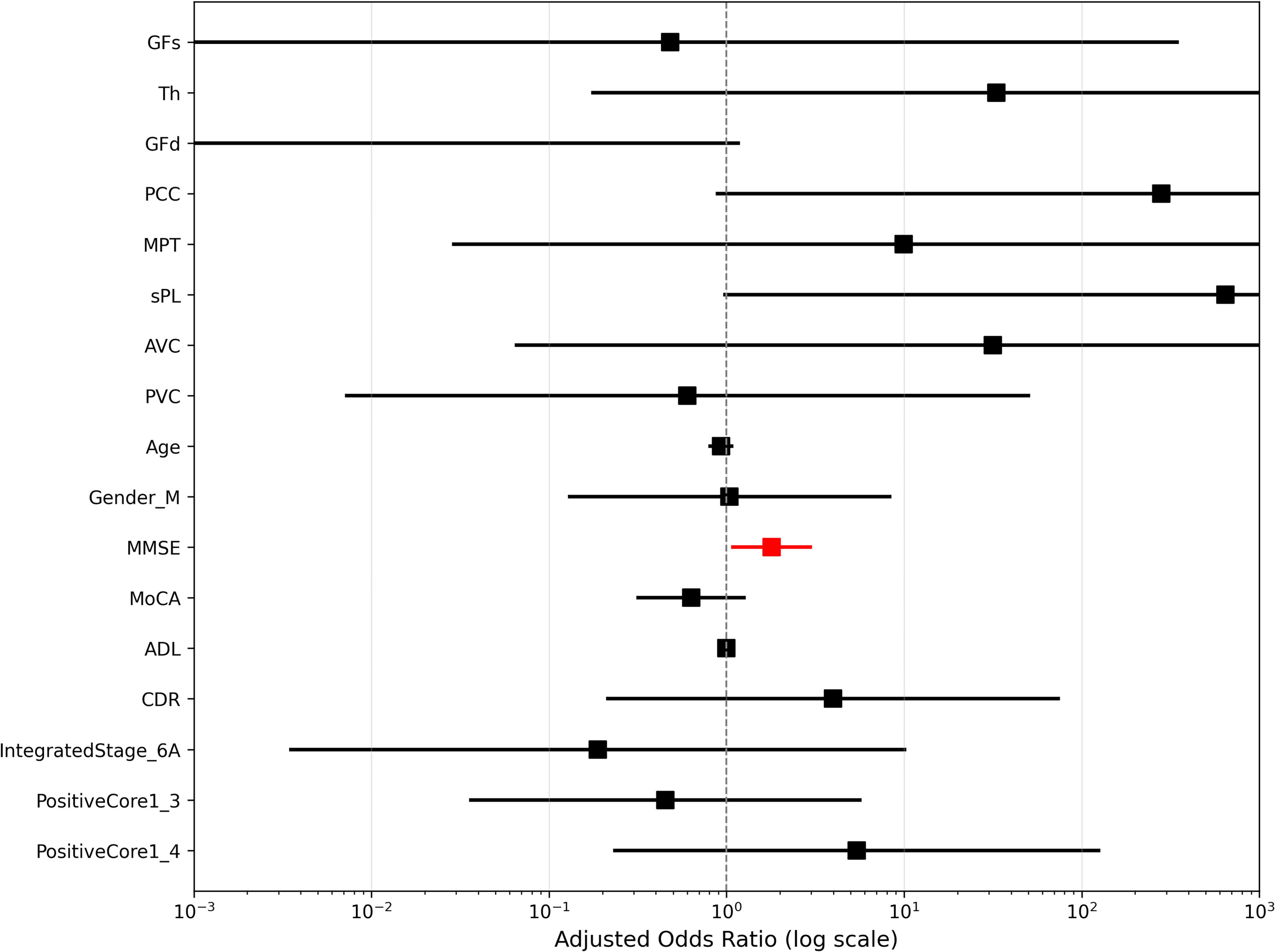
Forest plot of multivariable logistic regression. Adjusted odds ratios (ORs) from the multivariable logistic regression model after LASSO screening. Each square represents the adjusted OR for the corresponding variable, with horizontal lines indicating 95% confidence intervals. Red lines/squares denote variables with P < 0.05 (MMSE), while black denotes those with P ≥ 0.05. The model included the 8 DOI SUVRs indicators retained by LASSO and all covariates.

### Model Performance and Validation

The apparent AUC of the final model was 0.920, which decreased to an optimism-corrected AUC of 0.780 (95% CI: 0.708–0.884) after 1000-bootstrap resampling, with an optimism of 0.139, indicating moderate overfitting (Figure 4). Calibration was acceptable (Brier score = 0.097), although the Hosmer–Lemeshow test suggested possible calibration bias (χ² = 18.49, df = 8, P = 0.018; Supplementary data 1). DCA demonstrated some net clinical benefit within the threshold probability range of 0.2–0.7, though the overall gain was modest (Figure 5).

**Figure 4.**
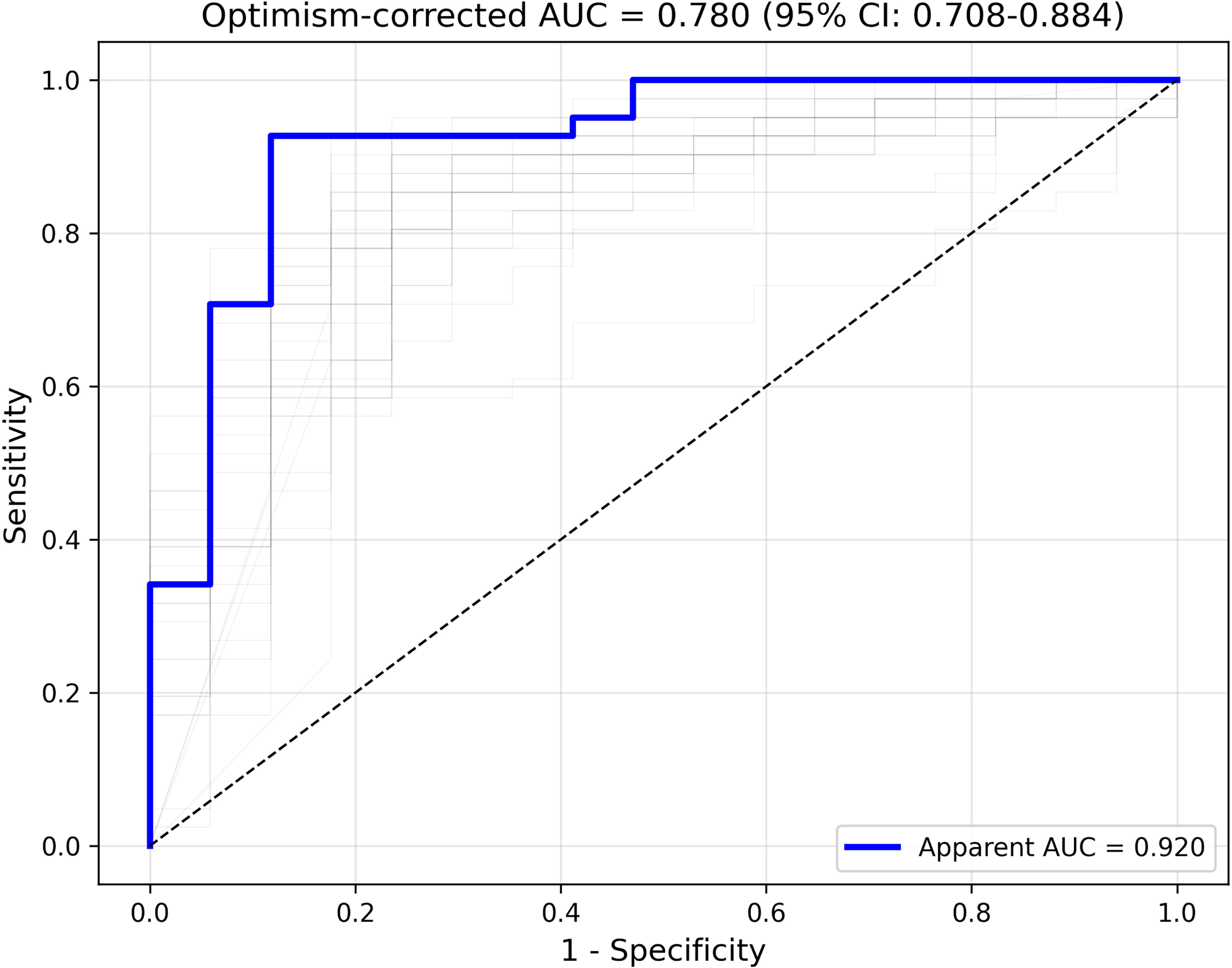
ROC curve of the final prediction model. The bold blue line represents the apparent ROC curve (Apparent AUC = 0.920). The 100 thin light□gray lines in the background are randomly selected ROC curves from 1,000 bootstrap resamples, illustrating the variability of the model across different resampled datasets. After 1,000 bootstrap corrections, the optimism□corrected AUC was 0.780 (95% CI: 0.708–0.884).

**Figure 5.**
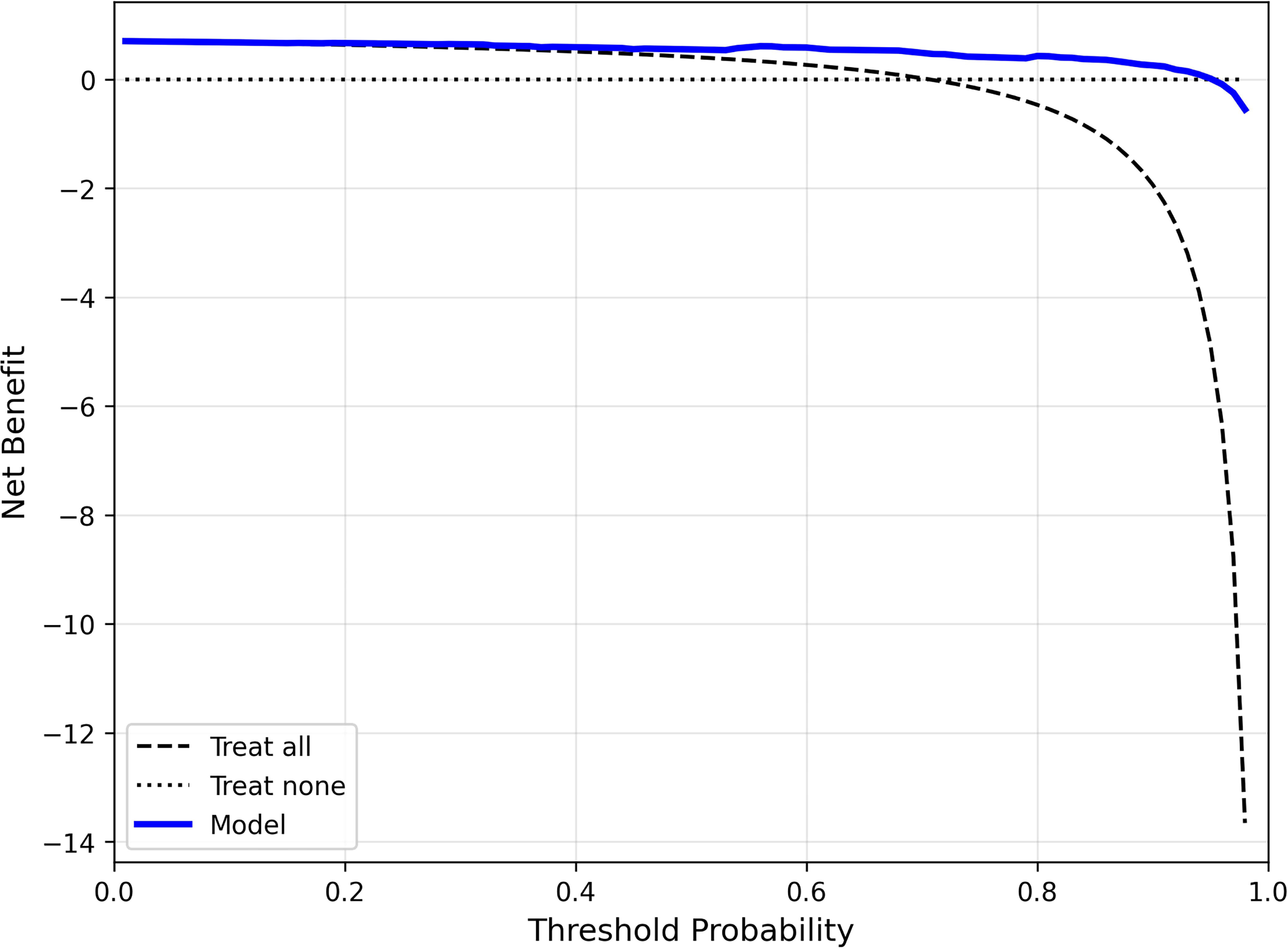
Decision curve analysis (DCA) for the final prediction model. The x□axis represents threshold probability, and the y□axis represents net benefit. The black dashed line indicates the net benefit of the “treat□all” strategy, while the black dotted line represents the “treat□none” strategy (net benefit = 0). The solid blue line shows the net benefit when decisions are guided by the model. Where the model curve lies above both extreme□strategy lines within a given threshold range, it suggests that using the model provides clinical net benefit within that range.

### Sensitivity Analyses

Backward stepwise regression (AIC-based) retained MMSE, ADL, and positive core 1 biomarkers 4, with AIC = 70.79 and AUC = 0.693—consistent with the LASSO model in preserving MMSE. In 100 random 2:1 splits, the mean test-set AUC was 0.74 ± 0.13 (range 0.40–0.87), reflecting model instability. When all ROI SUVRs were excluded, the covariates-only model yielded a Bootstrap-corrected AUC of only 0.574, confirming that PET DOI data provide substantial incremental predictive information beyond demographic and cognitive variables alone.

## Discussion

### LVA for AD: Current Status and Challenges

Single□center studies have identified several major obstacles to the widespread adoption of LVA for AD: the overall response rate is modest and varies considerably across centers (60–84.2%) [6–8]; clinical benefits tend to decline over time [8,13]; and the determinants of treatment response remain poorly understood, making it difficult to identify which patients are most likely to benefit. These uncertainties have fueled intense controversy in China, even though conservative estimates place the nationwide caseload at over a thousand applications [14–16]. To address these issues, the nationwide multi□center CLEAN□AD trial, conducted under a unified protocol, has recently been launched and is expected to provide more definitive evidence. Meanwhile, small□sample pilot trials are ongoing in the United States (REMIND) and Singapore (CLyVeB□AD□1), and the successful establishment of LVA animal models [17,18] offers a valuable platform for mechanistic exploration.

From a technical perspective, current LVA procedures mainly include: (1) lymphatic vessel to external jugular vein anastomosis; (2) lymphatic flap to internal jugular vein (IJV) anastomosis; and (3) lymph node to external/internal jugular vein anastomosis, along with various combinations thereof. In our practice, we utilize the JDN at level IIA and the lymphatic flap at level III as donors, and preferentially select primary tributaries of the IJV (e.g., the facial vein or lingual vein) as recipients. This design takes advantage of the diastolic suction effect from the right atrium while avoiding systolic hydrostatic pressure from the large□caliber IJV. We have termed this technique high□flow upper cervical lymphatic□venous anastomosis (HUCLA), aiming to achieve a more proximal and theoretically enhanced lymphatic shunt. The presumed mechanism of LVA is to facilitate the clearance of metabolic waste from the MLVs, through the cervical lymphatic tissues, and ultimately into the jugular vein. The MLVs, as the proximal segment of this drainage pathway, may therefore be critical for interpreting LVA’s efficacy and refining patient selection.

### MLVs and AD: Anatomical and Functional Basis

The existence of MLVs was definitively established in 2015 by two landmark studies that provided both molecular and functional evidence [9,19]. These vessels are situated in the dura mater, tracking alongside the dural venous sinuses [9,20], and express all classical lymphatic endothelial cell markers [9,21]. Using high-resolution MRI, Absinta et al. visualized the MLVs in healthy volunteers and common marmosets, confirming that they course parallel to the superior sagittal and straight sinuses, as well as along branches of the middle meningeal artery, with diameters ranging from 7 to 842 µm [10].

Functionally, MLVs serve as a major conduit for draining CSF and its solutes into the peripheral lymphatic system. Tracers injected into the cisterna magna or brain parenchyma rapidly appear in the MLVs and subsequently accumulate in the deep cervical lymph nodes [9,22]. Conversely, ablation of MLVs dramatically reduces tracer accumulation in the lymph nodes and impairs brain-wide glymphatic CSF–interstitial fluid exchange [22–24], underscoring the essential role of MLVs in central nervous system waste clearance.

In mouse models of AD, ablation of MLVs not only exacerbates cerebral Aβ pathology but also induces Aβ deposition within the meninges—a phenomenon also observed in the dura of human AD patients [23]. Conversely, VEGF-C-mediated enhancement of MLV function has been shown to reduce Aβ burden and improve cognitive performance [23]. Together, these findings establish MLVs not only as a pathological nexus in AD progression but also as a promising therapeutic target, providing a biological rationale for surgical interventions—such as LVA—aimed at augmenting this drainage pathway.

### Quantification of A***β*** burden on PET

In AD PET imaging, SUVR remains the most widely used semi□quantitative metric, but its poor comparability across studies and tracers is a major limitation; using native□space FreeSurfer parcellation has been shown to improve concordance with histopathology [25]. The Centiloid scale was developed to harmonize SUVR values by linear transformation to a common 0–100 scale anchored to young controls and typical AD patients, thereby facilitating cross□center and cross□tracer comparisons [26]; however, it does not eliminate the inherent variability of the underlying quantification method, as evidenced by method□dependent amyloid positivity thresholds that can vary from 5.7 to 11.9 CL [27]. In contrast, Z□score analysis offers a statistical deviation□from□normality perspective by comparing individual regional or voxel□wise values to a normative database, which is particularly valuable for FDG PET pattern recognition and for detecting early focal changes before global metrics become abnormal [28,29].

Given the exploratory nature of this study and the practical constraints of a single□center design, we elected to use SUVR as our primary quantitative metric. SUVR is the most established and widely applied method in amyloid PET, enabling direct comparison with previous studies [30]. Additionally, our proposed scoring system intrinsically relies on regional SUVR values, and the available sample size did not permit robust Centiloid cross□calibration or the construction of a site□specific normative database for Z□score analysis. Thus, SUVR was considered a pragmatic and appropriate choice for this proof□of□concept investigation.

### The Design Rationale of the Novel Scoring System

The scoring system is fundamentally grounded in the hydrodynamic principles of the LVA shunt system. As previously described, the identified human MLVs are predominantly located along the SSS and SS, running parallel to them. Accordingly, we assign higher anatomical scores to ROIs closer to these sinuses, based on the premise that proximity to MLVs facilitates more efficient clearance. Although global Aβ burden alone does not correlate directly with cognitive performance in AD, the ratio of cortical to subcortical Aβ deposition has been shown to associate significantly with semantic verbal fluency [31], suggesting that regional Aβ distribution may carry clinical relevance. We therefore also assign functional scores to ROIs theoretically implicated in AD symptomatology. These anatomical and functional scores are then integrated through a weighted composite system. The observation that the univariately significant predictor S_4+5_ encompassed sPL and AVC (adjacent to the mid□posterior SSS) and PCC (adjacent to the anterior SS) provides empirical support for the anatomical logic of this scoring framework.

### Limitations

Several limitations should be acknowledged. First, this was a single-center retrospective study with a small sample (n=58), limiting generalizability and statistical power, and the moderate optimism (0.139) in the corrected AUC suggests some overfitting. Second, the scoring system relied on anatomical proximity to the SSS/SS rather than direct MLV visualization, potentially missing individual variations in MLV distribution. Third, rigid registration using NeuroQ software may introduce misalignment errors, particularly in atrophic brains. Fourth, SUVR values are protocol-dependent and not directly comparable across centers or tracers without cross-calibration. Fifth, regional Aβ deposition may be confounded by APOE ε4 status [32], age-related comorbidities, and cerebral amyloid angiopathy [33], which were not accounted for. Sixth, unmeasured factors such as baseline glymphatic function or intraoperative variables may have influenced outcomes. Finally, post-selection inference from LASSO may underestimate standard errors, and the wide CIs for adjusted ORs reflect estimate instability. These findings should therefore be interpreted as hypothesis-generating and require validation in larger, prospective multicenter studies.

Despite these limitations, this work represents a first step toward imaging-guided patient selection for LVA. The scoring system’s design—grounded in MLV anatomy and hydrodynamic principles—offers a mechanistically informed alternative to global Aβ metrics. Future studies should validate this system in larger, multicenter prospective cohorts, incorporate longitudinal PET data, and explore multimodal fusion with MRI structural connectivity, CSF biomarkers, and gene risk profiles.

### Conclusion

This exploratory study introduces a novel Aβ PET scoring system that quantifies regional amyloid burden according to proximity to MLVs, and demonstrates predictive potential for LVA response in AD when combined with baseline MMSE. The SUVR of DOIs provided substantial incremental value over demographic and cognitive covariates alone. These findings require confirmation in larger, multicenter prospective studies with external validation before clinical translation.

## Data Availability

All data produced in the present study are available upon reasonable request to the authors.

